# Automation Bias in AI-Assisted Detection of Cerebral Aneurysms on Time-of-Flight MR-Angiography

**DOI:** 10.1101/2024.05.31.24308021

**Authors:** Su Hwan Kim, Severin Schramm, Evamaria Olga Riedel, Lena Schmitzer, Enrike Rosenkranz, Olivia Kertels, Jannis Bodden, Karolin Paprottka, Dominik Sepp, Martin Renz, Jan Kirschke, Thomas Baum, Christian Maegerlein, Tobias Boeckh-Behrens, Claus Zimmer, Benedikt Wiestler, Dennis M. Hedderich

## Abstract

**Background:** AI systems have the potential to support in detecting cerebral aneurysms. Yet, the role of automation bias (inclination of humans to overly rely on automated decision-making systems) in AI-assisted cerebral aneurysm detection remains unclear.

**Purpose:** To determine how automation bias can affect radiologists with varying experience levels when reading time-of-flight magnetic resonance angiography (TOF-MRA) studies with the assistance of an AI system for cerebral aneurysm detection.

**Methods:** In this prospective experiment, nine radiologists with varying levels of experience evaluated twenty TOF-MRA exams for the presence of anterior circulation aneurysms, with each arterial segment rated on a 4-point Likert scale, and provided follow-up recommendations. Every case was evaluated twice (with or without assistance by the AI software © mdbrain), with a washout-period of at least four weeks between the two sessions. Ten out of twenty cases included at least one false-positive AI finding. Aneurysm ratings, follow-up recommendations, and reading times were assessed using the Wilcoxon signed-rank test. A thematic analysis was performed to summarize reader feedback and observations.

**Results:** False-positive AI results led to significantly higher suspicion of aneurysm findings (p = 0.01). Inexperienced readers further recommended significantly more aggressive follow-up examinations when presented with false-positive AI findings (p = 0.005). Reading times were significantly shorter with AI assistance in inexperienced (164.1 vs 228.2 seconds; p < 0.001), moderately experienced (126.2 vs 156.5 seconds; p < 0.009), and very experienced (117.9 vs 153.5 seconds; p < 0.001) readers alike.

**Conclusion:** Our results demonstrate susceptibility of radiology readers to automation bias in detecting cerebral aneurysms in TOF-MRA studies when encountering false-positive AI findings. In inexperienced readers, this behavior further translated into more aggressive follow-up recommendations. AI assistance resulted in significantly shorter reading times across experience levels. While AI systems for cerebral aneurysm detection can provide benefits, challenges in human-AI interaction need to be mitigated to ensure safe and effective adoption.

## Introduction

Cerebral aneurysms have an estimated prevalence of 2% and account for up to 85% of non-traumatic subarachnoid hemorrhages (SAH), which are associated with a considerable risk of severe disability and mortality (1). Early detection of aneurysms allows for timely rupture risk assessment and optimal management, potentially enhancing patient outcomes (2). While digital subtraction angiography (DSA) remains the gold standard imaging modality, computed tomography angiography (CTA) and time-of-flight magnetic resonance angiography (TOF-MRA) have proven to be reliable non-invasive methods for detecting cerebral aneurysms (3,4). Improvements of these diagnostic imaging techniques have further led to an increased detection of unruptured cerebral aneurysms over time (5,6).

In recent years, numerous studies have explored the use of artificial intelligence (AI) computer-assisted diagnosis (CAD) systems for the detection of cerebral aneurysms in CTA, MRA or DSA datasets (7). These included both studies performing a standalone evaluation of an AI CAD system (8–13) and ones applying AI CAD systems as a reader-aid (14–18).

Yet, how cognitive biases affect the diagnostic performance of radiologists when interacting with AI CAD systems for cerebral aneurysm detection remains unknown. One such phenomenon that has been described in the context of AI-assisted diagnosis is automation bias, which is the tendency of humans to overly rely on automated decision-making systems (19–23). For instance, one recent study demonstrated that radiologists are prone to favor even incorrect suggestions from an AI-based mammogram classification system (22). Notably, automation bias is a major concern in high-sensitivity contexts, such as AI-based cerebral aneurysm detection, where a high frequency of false positives is expected (7).

The aim of this study was to determine how automation bias can affect radiologists with varying experience levels when reading TOF-MRA studies with the assistance of an AI system for cerebral aneurysm detection.

## Methods

Institutional review board approval was obtained, and the need for informed consent was waived.

### Dataset and AI CAD System

The dataset consisted of a total of twenty 3D TOF-MRA studies acquired between 06/2021 and 12/2023 at a collaborating outpatient radiology practice (“Die Radiologie”, Munich, Germany) at which a CAD AI system (© mdbrain, version 4; Mediaire GmbH) for cerebral aneurysm detection is routinely used. TOF-MRA images with annotations of AI-detected findings were automatically generated by the software. Details regarding the training of the AI model have been reported previously (24). Image studies had been acquired using two clinical 3T scanners (Skyra, Siemens Healthineers, Erlangen, Germany; Ingenia Elition X, Philips Healthcare, Best, The Netherlands) and two 1.5 T scanners (Magnetom Aera, Siemens Healthineers, Erlangen, Germany; Ingenia, Philips, Best, Netherlands). Local routine protocols were applied, with the slice thickness ranging between 0.6 – 1.4 mm.

To identify both cases with true-positive and false-positive AI findings, a retrospective full-text search was performed within the radiology information system (RIS). The final dataset included ten cases with at least one false-positive AI finding and ten cases with at least one true-positive AI finding but no incorrect AI findings. In total, the dataset included twelve aneurysms (eleven saccular and one fusiform aneurysm). Only findings of the anterior circulation were considered for the purpose of this study. Cases were selected and verified by a board-certified interventional neuroradiologist.

TOF-MRA studies were independently reviewed by two senior neuroradiologists (16 years and 9 years of neuroradiology experience each) for the presence of cerebral aneurysms. Full consensus was reached between the two neuroradiologists in all twenty cases. Digital subtraction angiography (DSA) was only available in a single case. False-positive AI findings were classified as vascular loop (5/10), infundibulum (3/10), or perforator (2/10) (Supplement 1).

### Reader Study

Anonymized image datasets with and without AI annotations were imported to our local Picture Archiving and Communicating System (PACS) system (IDS7, Sectra Medical Systems AB, USA). Cases were randomized into two sets (A and B), each containing four and six false-positive cases.

A total of nine readers evaluated the dataset twice, with a washout-period of at least four weeks between the two sessions (average: 45 days) (Figure 1). Participants included three inexperienced (radiology residents with 6-12 months of neuroradiology experience), three moderately experienced (board-certified radiologists), and three very experienced readers (board-certified neuroradiologists) (Table 1). In each session, one set of cases was evaluated with AI assistance and the other without. Before the first session, five sample cases with verified true-positive AI findings were showcased to familiarize the readers with the AI annotations and cultivate trust in the tool’s accuracy. To define a clear interaction protocol, readers were instructed to examine AI annotations before reviewing the original TOF-MRA image series (AI as “first reader”). By default, only the axial TOF-MRA series (with and without AI annotations) were included in the hanging protocol, but readers were allowed to perform multiplanar or 3D reconstructions at will.

**Figure 1:**
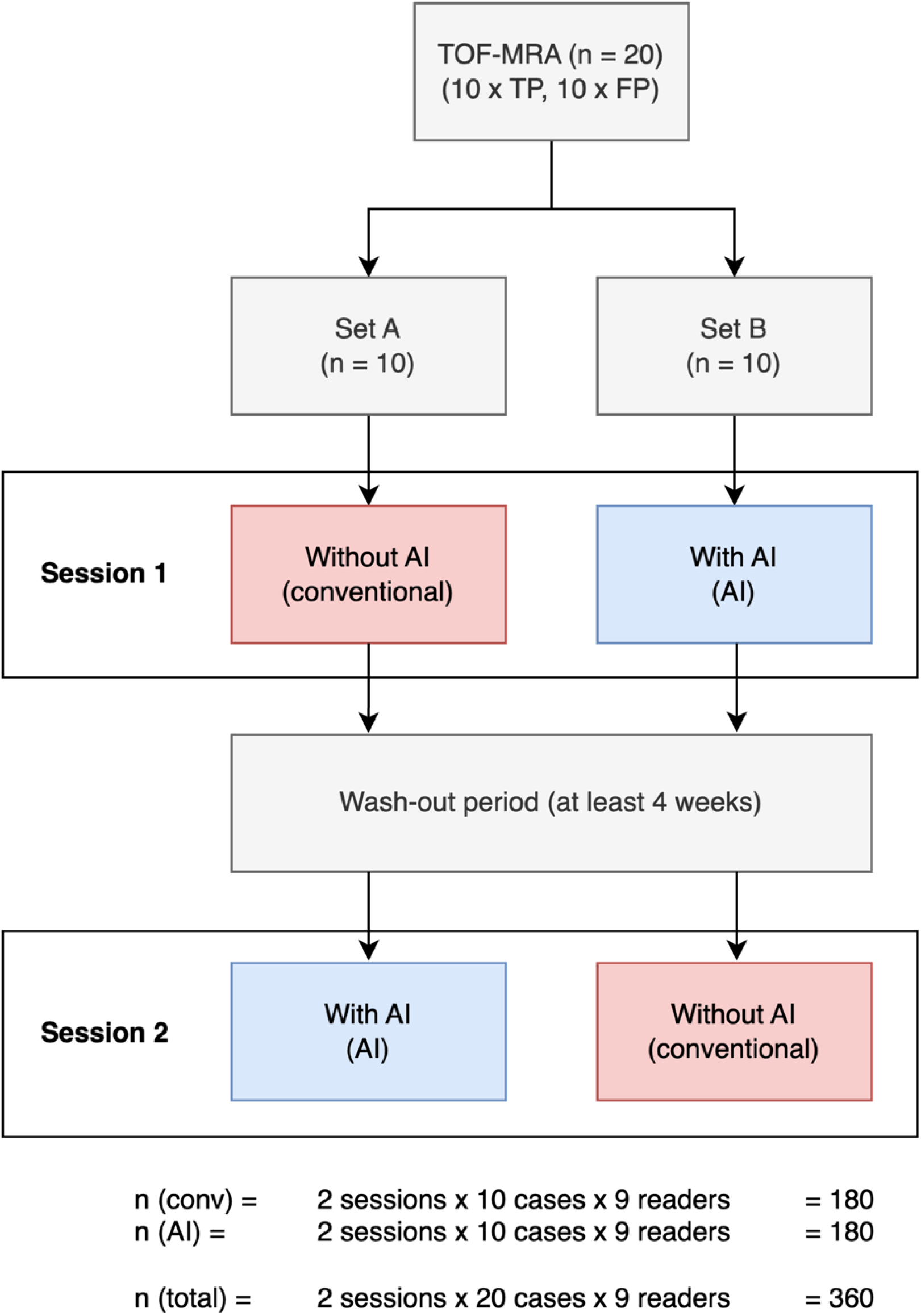
Study Design. A total of nine readers evaluated the dataset twice, with a washout-period of at least four weeks between the two sessions. In each session, one set of cases was evaluated with AI assistance and the other without.

**Table 1:** Overview of readers.

| **reader_id** | **Experience Level** | **Age Range** | **Sex** | **Radiology Experience (in years)** | **Neuroradiology Experience (in years)** | **Experience in Neuroradiology Interventions (in years)** |
| --- | --- | --- | --- | --- | --- | --- |
| **E1R1** | Inexperienced | 26 – 30 | Female | 1 | 1 | 0 |
| **E1R2** | Inexperienced | 26 – 30 | Female | 0.5 | 0.5 | 0 |
| **E1R3** | Inexperienced | 26 – 30 | Female | 0.5 | 0.5 | 0 |
| **E2R1** | Moderately experienced | 31 – 35 | Female | 6.5 | 2.5 | 2 |
| **E2R2** | Moderately experienced | 31 – 35 | Male | 6 | 2 | 1 |
| **E2R3** | Moderately experienced | 36 – 40 | Female | 5 | 3 | 1 |
| **E3R1** | Very experienced | 41 – 45 | Male | 8 | 6 | 4 |
| **E3R2** | Very experienced | 36 - 40 | Male | 9 | 4 | 3 |
| **E3R3** | Very experienced | 46 - 50 | Male | 18 | 14 | 10 |

For each arterial segment of the anterior circulation, readers provided a 4-point Likert-scale rating on the presence of a cerebral aneurysm (1: certainly not present, 2: likely not present, 3: likely present, 4: certainly present). In addition, for each aneurysm (not each patient), readers indicated a follow-up recommendation (no follow-up examination, MRI, or DSA). The patient age was provided only upon request. Aneurysm ratings and recommendations were documented using an online form tool (© Google Forms, Google Inc., Mountain View, USA) and reading times were recorded using a time tracking software (Toggl Track, © Toggl OÜ, Tallinn, Estonia).

### Analysis

Data manipulation, data visualization, and statistical analyses were performed using Python (version 3.9.7).

To prevent the misclassification of findings due to inaccurate locations, findings with a Likert scale rating of 2 or more were grouped as ‘Acom (anterior communicating artery)’ if the location was described as ‘A1 segment (anterior cerebral artery)’, ‘Acom’, or ‘A2 segment (anterior cerebral artery)’, and as ‘ICA (internal carotid artery)’ if the location was described as ‘ICA’ or ‘terminal T’. Follow-up recommendations were modeled as an ordinal scale from 0 to 2 (0: no follow-up, 1: MRI, 2: DSA), reflecting the level of invasiveness. The degree of bias towards false-positive AI findings was quantified by comparing aneurysm ratings and follow-up recommendations for false-positive AI findings between the two reading workflows (conventional vs with AI).

Normality of data distribution was evaluated using the Shapiro-Wilk test. The level of statistical significance was set at p = 0.05. Accounting for the paired nature of the data (rating of the same findings with or without AI), a Wilcoxon signed-rank test was used to evaluate statistical significance for aneurysm ratings, follow-up recommendations, and reading times.

Sensitivity (per lesion), sensitivity (per patient) and specificity (per patient) are reported, grouped by reading workflows and experience levels. Binary classifications for the presence of aneurysms were inferred from the 4-point Likert scale ratings (1-2: absent, 3-4: present). The diagnostic classification per patient was performed as described previously (18): A case was classified as true positive only when the reader identified all aneurysms without reporting any false positives. True negatives were defined as cases where both the reader and the reference standard agreed on the absence of any aneurysms. Cases with at least one false positive finding were labeled as false positive. Cases where the reader failed to detect a true aneurysm were considered false negatives.

To illustrate the impact of automation bias, rating pairs for false-positive AI findings (conventional vs with AI) were visualized using Sankey diagrams, grouped by experience level. For individual readers, associations between diagnostic performance metrics, mean reading times, and frequencies of unconfident Likert scale ratings (2 or 3) were determined using the Spearman’s correlation coefficient. Results were displayed in a color-coded correlation matrix. A thematic analysis was performed to summarize reader feedback and observations.

## Results

### Automation Bias

Exposure of readers with false-positive AI recommendations led to significantly higher Likert scale ratings of suspected aneurysms (p = 0.01). Ratings of inexperienced readers were particularly influenced by false-positive AI findings (p = 0.002), whereas no significant differences were observed in the subgroup analyses of moderately experienced (p = 0.18) and very experienced readers (p = 0.59). Most instances where aneurysm ratings increased with AI involved arterial segments that were confidently rated as unremarkable (rating of 1) without AI support (Figure 2).

**Figure 2:**
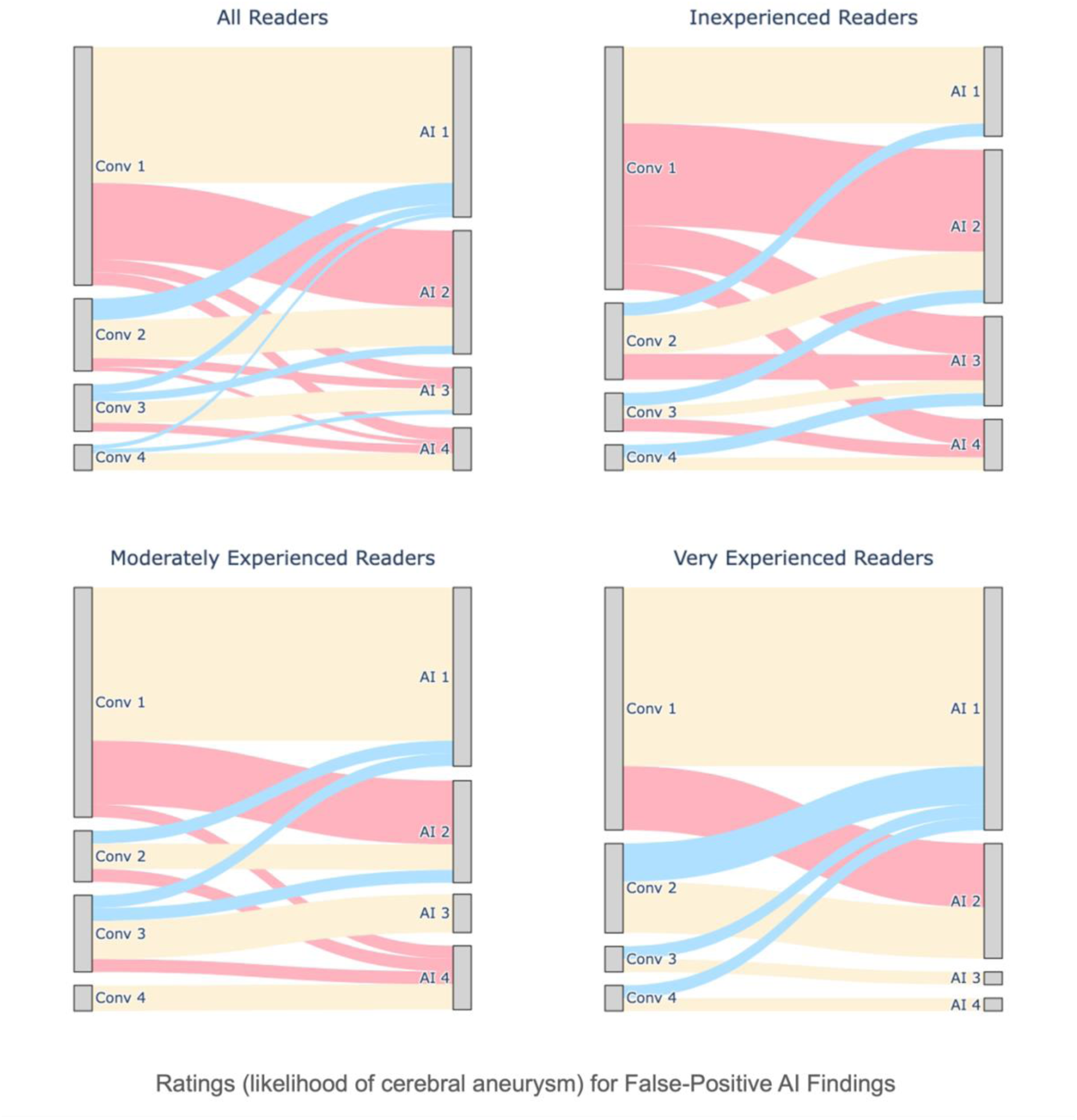
Likert scale ratings (by workflow and experience level) for false-positive AI findings. The Sankey diagrams illustrate how readers rated identical vascular segments for the presence of aneurysms in the presence (‘AI’) and absence (‘Conv’) of false-positive AI results on a Likert scale from 1-4 (1: certainly absent, 2: likely absent, 3: likely present, 4: certainly present). For instance, ‘Conv 3’ indicates that a vascular segment was rated as ‘aneurysm likely present’ without AI assistance. Connections from left to right nodes are marked red, yellow, or blue depending on whether ratings were higher, identical, or lower with AI assistance than without (conventional). Inexperienced readers were early residents with less than 1 year of neuroradiology experience, moderately experienced readers board-certified radiologists, and very experienced readers certified neuroradiologists.

### Follow-up recommendations

Follow-up recommendations for false-positive AI findings varied by reader experience (Figure 3). Overall, readers more frequently recommended MRI and DSA in the AI-assisted workflow (conventional: MRI = 24, DSA = 8; AI: MRI = 36, DSA = 7), although the difference was not significant (p = 0.21). Inexperienced readers recommended significantly more invasive follow-up examinations (p = 0.005), with a marked increase in MRI recommendations with AI (conventional: MRI = 9; AI: MRI = 21) but consistent DSA recommendations (1 in both workflows). Moderately experienced readers showed a slight increase in MRI recommendations (conventional: 7; with AI: 9) and a minimal reduction in DSA recommendations (conventional: 4, with AI: 3) with AI (p = 1.0). Likewise, very experienced readers displayed only minimal changes in follow-up patterns across workflows (conventional: MRI = 8, DSA = 3; with AI: MRI = 6, DSA = 3) (p = 0.71).

**Figure 3:**
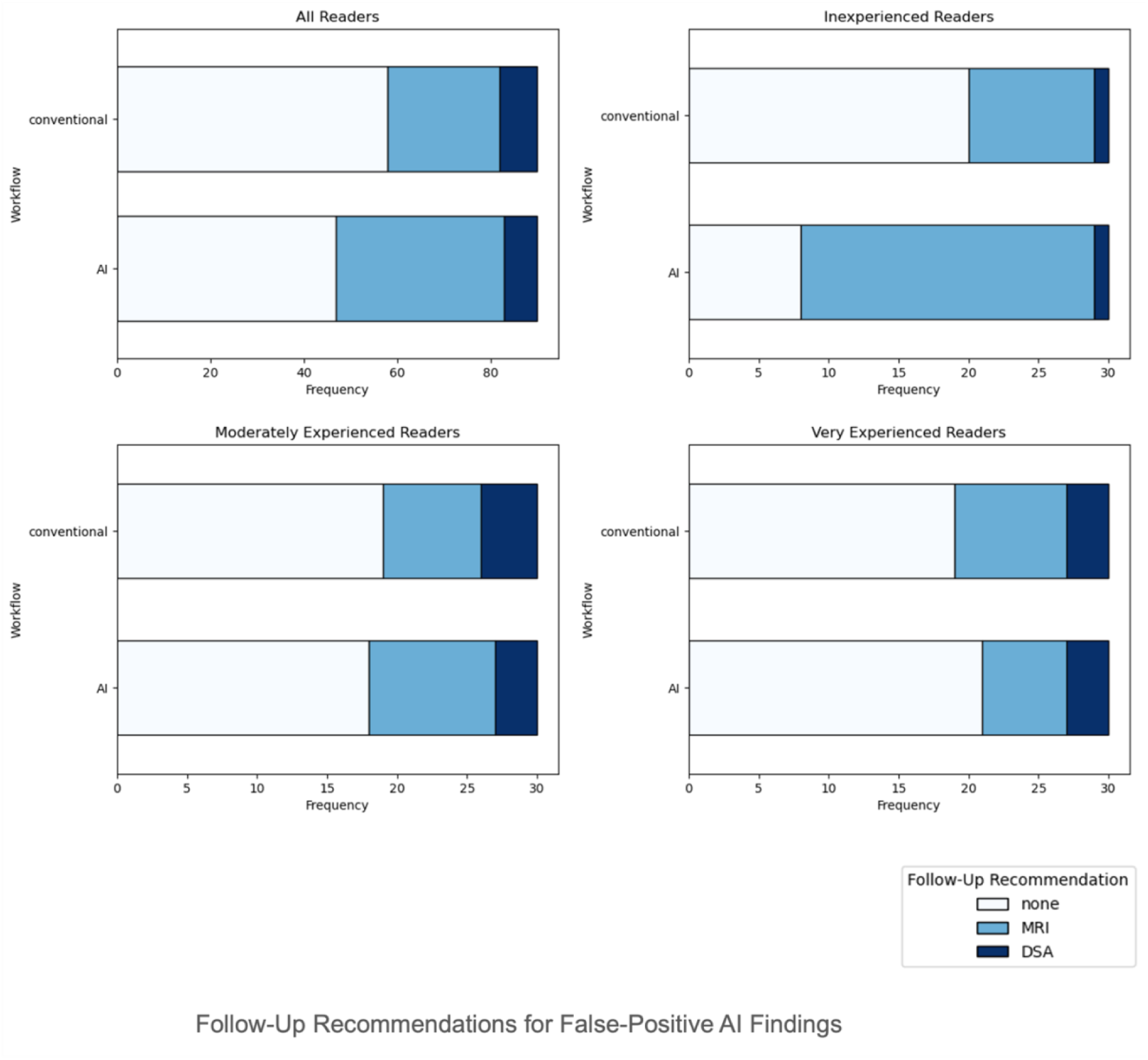
Follow-up recommendations (by workflow and experience level) for false-positive AI findings. Recommendations were modeled as an ordinal scale from 0 to 2 (0: no follow-up, 1: follow-up MRI, 2: digital subtraction angiography; DSA), reflecting the level of invasiveness. Significantly more invasive follow-up recommendations were observed with AI assistance in inexperienced readers (p = 0.005), but not in moderately experienced (p = 1.0) or very experienced readers (p = 0.71).

### Interpretation times

Mean reading times were significantly shorter with AI assistance in inexperienced (164.1 vs 228.2 seconds; p < 0.001), moderately experienced (126.2 vs 156.5 seconds; p < 0.009), and very experienced (117.9 vs 153.5 seconds; p < 0.001) readers alike. Overall, mean reading times were reduced significantly from 179.4 to 136.0 seconds with AI support (p << 0.001) (Figure 4).

**Figure 4:**
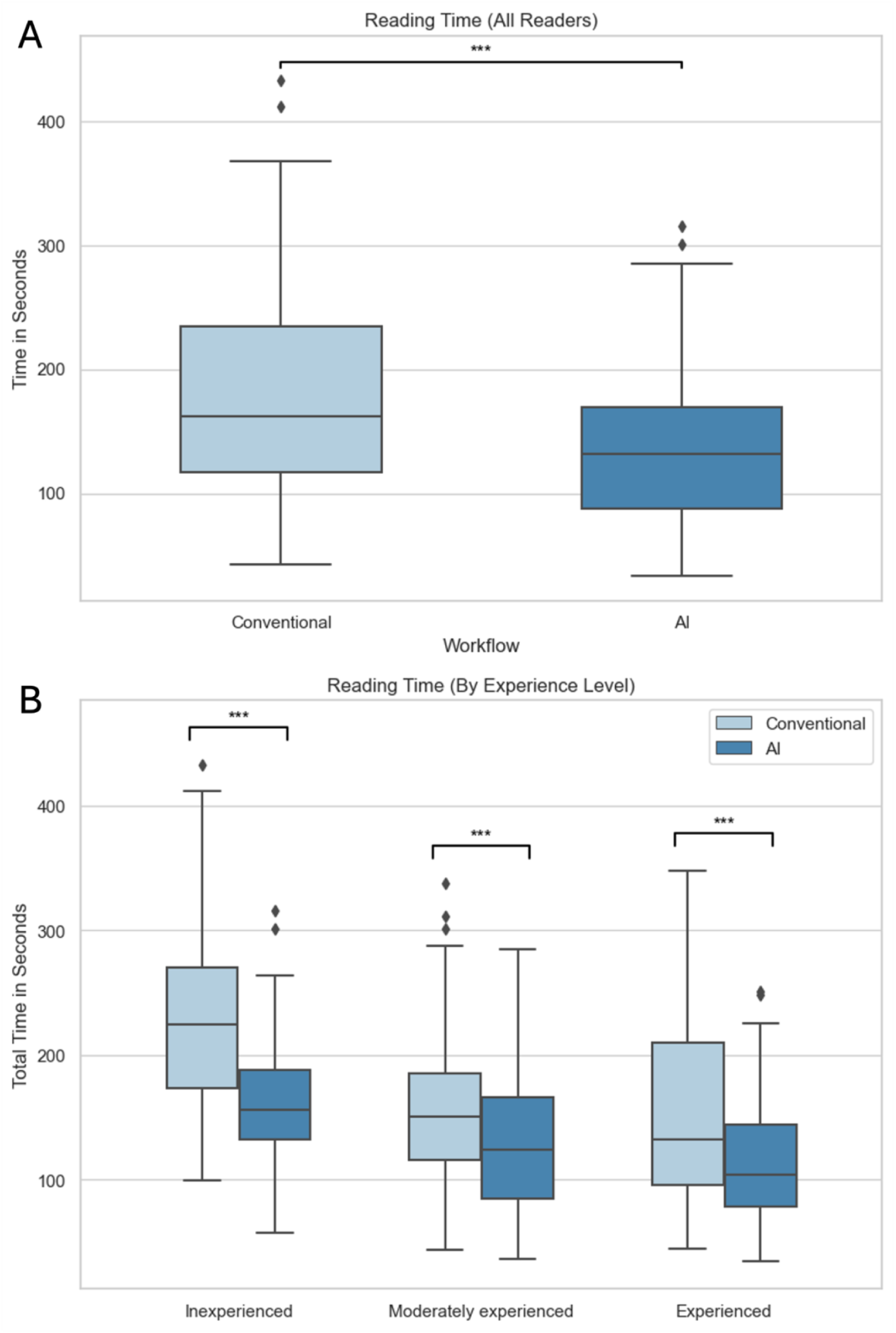
Reading times. A: Overall. B: By experience level.

### Diagnostic Performance

Overall, the combined per-lesion and per-patient sensitivity both increased from 0.88 in the conventional workflow to 0.97 with AI assistance. Specificity per patient remained unchanged at 0.79 in both workflows (Table 2). Inexperienced readers exhibited a marked improvement in per-lesion and per-patient sensitivity with AI (from 0.69 to 1.00 for both), which, however, came at the cost of decreased specificity (conventional: 0.81, with AI: 0.66). Moderately experienced readers showed exceptional per-lesion and per-patient sensitivity in both workflows (consistently at 0.97) and reached higher specificity with AI assistance (conventional: 0.70, with AI: 0.79). Similarly, very experienced readers displayed very high sensitivity on lesion level (conventional: 0.94, with AI: 0.97) and patient level (conventional: 0.97, with AI: 0.94), while also demonstrating superior specificity over the other reader groups (conventional: 0.88, with AI: 0.94).

**Table 2:** Diagnostic performance by reader group and workflow (all cases).

| Reader Group | Sensitivity (per lesion) |  | Sensitivity (per patient) |  | Specificity (per patient) |  |
| --- | --- | --- | --- | --- | --- | --- |
|  | Conventional | With AI | Conventional | With AI | Conventional | With AI |
| **Inexperienced**<br>**(n = 3)** | 0.69 | 1.00 | 0.69 | 1.00 | 0.81 | 0.66 |
| **Moderately experienced**<br>**(n = 3)** | 0.97 | 0.97 | 0.97 | 0.97 | 0.70 | 0.79 |
| **Very experienced**<br>**(n = 3)** | 0.94 | 0.97 | 0.97 | 0.94 | 0.88 | 0.94 |
| **Overall**<br>**(n = 9)** | 0.88 | 0.97 | 0.88 | 0.97 | 0.79 | 0.79 |

### Feedback and Observations

Qualitative findings obtained from reader feedback and observations were summarized in Table 3. There was overwhelming agreement that high sensitivity of the AI tool was far more important than specificity, given that the consequences of missing a true aneurysm are more severe. At the same time, many readers trusted that the commercial AI software must have been optimized for high sensitivity and showed little concern about aneurysms potentially missed by the AI. Most readers appreciated the AI’s utility in filtering out potentially relevant findings that require more careful review by the human reader. The AI assistance was perceived as reassuring, particularly by the inexperienced readers.

**Table 3:** Reader feedback and observations.

| Theme | Feedback and Observations |
| --- | --- |
| **Diagnostic performance** | - High sensitivity is more important than specificity, given that consequences of missing an aneurysm could be critical<br>- Many readers trusted that AI must have been optimized for high sensitivity |
| **Clinical utility and adoption** | - Human readers should sort out false-positive findings<br>- Particularly useful for distal findings easy to overlook<br>- Helps to direct attention to potentially relevant findings<br>- False-positive AI findings may artificially create morbidity leading to invasive procedures with potential complications (e.g. DSA) |
| **Psychological aspects** | - It is reassuring to have an AI double-check findings<br>- Due to medicolegal implications, it is difficult to not follow-up on AI-positive findings, even when an aneurysm is considered very unlikely<br>- The availability of AI results promotes complacency, radiologists should make a deliberate effort to systematically review all arterial segments even in the presence of an AI assistance<br>- Clinical referrers are unsettled when AI-positive findings are reported, radiologists are required to justify contradicting opinions<br>- In the presence of one aneurysm, very experienced radiologists tend to carefully look for the presence of further aneurysms (opposite of “satisfaction of search”) |

However, readers remarked that the availability of an automated aneurysm detection software promotes complacency, and that radiologists should make a deliberate effort to review the images systematically, nonetheless. Interestingly, two radiologists stated that they would feel compelled to follow-up on AI-positive findings due to potential medicolegal repercussions, even when they personally doubted the presence of an aneurysm. One reader noted that referring physicians are equally unsettled when made aware of AI-positive findings, necessitating that radiologists justify any opposing assessments. On a 5-point Likert scale (1: not at all helpful, 5: very helpful), the AI software received a median rating of 4.

### Reader-Level Correlation

Correlation of reader-specific metrics reveals important associations (Figure 5). Notably, the level of neuroradiology experience showed a strong positive correlation with per-patient accuracy and a moderate positive correlation with specificity. Experience negatively correlated with mean reading time and number of unconfident ratings (Likert scale rating of 2 or 3), indicating that experienced readers were faster and had more confidence in their assessments. The number of unconfident ratings positively correlated with mean reading time and negatively correlated with patient-level accuracy, possibly mediated by experience level.

**Figure 5:**
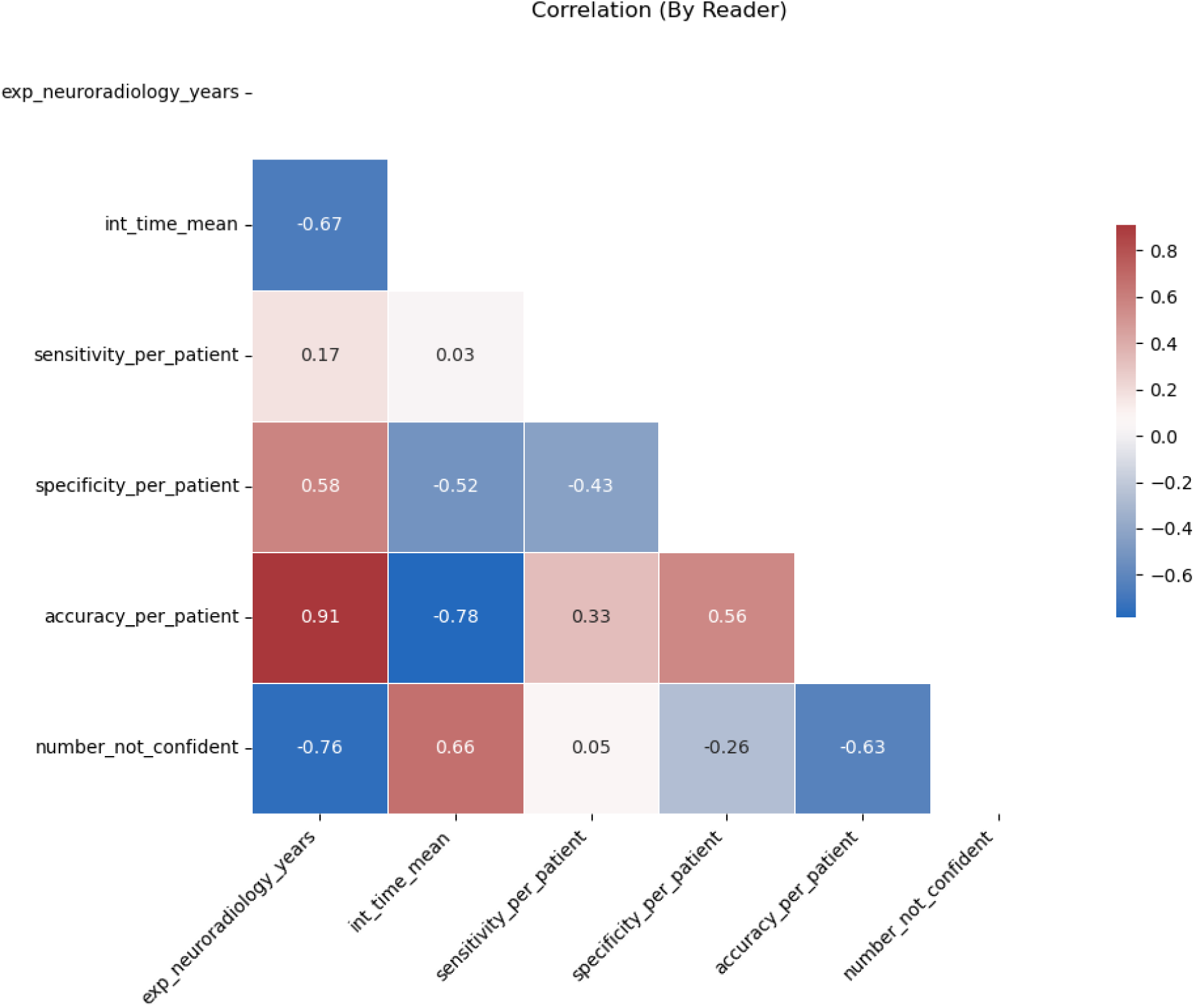
Correlation matrix (by individual reader). The Spearman’s correlation coefficient was calculated to determine the association between diagnostic performance metrics, mean reading times, and frequency of unconfident ratings (Likert scale rating of 2 or 3).

## Discussion

This study explored how automation bias can affect radiologists with varying experience levels when evaluating TOF-MRA studies for the presence of cerebral aneurysms with the assistance of a CAD AI software.

Radiologists were more likely to believe the presence of an actual aneurysm when presented with false-positive AI findings, demonstrating their susceptibility to automation bias. This observation corroborates previous warnings about the adverse effects of over-reliance on AI tools in medical imaging (25–27). While inexperienced readers were strongly influenced by the AI findings, very experienced readers demonstrated resilience to this bias. These results are consistent with an earlier study on mammography reading revealing diminishing degrees of automation bias with greater reader experience (22), underlining the critical role of experienced radiologists in validating AI results.

Importantly, inexperienced readers were also significantly more likely to recommend follow-up exams in response to false-positive AI findings. This indicates that incorrect AI suggestions can engender unnecessary procedures, raising both costs and patient anxiety. Future studies should explore the health economic implications of such redundant medical services caused by erroneous AI findings. Intriguingly, some of the more experienced readers commented that in actual practice, they might choose to recommend follow-up examinations for AI-positive findings even when an aneurysm appeared improbable, simply to avoid medicolegal risks. Extending earlier concerns about legal liability for errors committed by AI systems (28,29), this finding suggests that fears of legal repercussions for overriding correct AI results can lead to medically irrational decisions. These concerns are not fully unfounded, considering that certified AI systems in radiology commonly have intended-use statements with disclaimers passing the medicolegal responsibility onto the user (30). Clear guidelines and legislation are necessary to empower physicians to make decisions based on medical expertise and patient needs, rather than legal concerns.

One practical challenge pointed out by several readers was the lack of standardized guidelines for the management of cerebral aneurysms. According to the latest 2022 *European Stroke Organisation (ESO)* guidelines, follow-up imaging frequency and duration should be determined “*based on aneurysm– and patient-related risk factors for growth or rupture, and risk of treatment”* (31). Although this statement reflects the complexity of aneurysm management and the individuality of each patient case, it does not offer practical guidance for radiologists in establishing a consistent follow-up strategy for incidental aneurysms. A 2018 survey study revealed considerable heterogeneity among neuroradiologists in follow-up recommendations for small (< 7 mm) unruptured aneurysms, further underscoring the necessity for standardized protocols (32).

Unlike a previous study that reported mixed effects of AI assistance on TOF-MRA reading times of three radiology readers and three students (18), this study observed decreased reading times across all experience levels. The fact that this efficiency improvement was demonstrated in an artificial cohort with an extraordinarily high rate of false-positive AI findings (50%) is noteworthy, since false-positive findings have been suspected to cause increased workload (7).

### Limitations

This study has several limitations.

First, the reference standard was defined by expert consensus rather than DSA, which is widely regarded as the gold standard modality, but was available in only one case. Nonetheless, the fact that the two experts who defined the reference standard and the most senior reader participating in the study (14 years of neuroradiology experience) had a 100% agreement (without AI), suggests a robust reference standard.

Second, the readings took place in a controlled study setting and readers might behave differently in clinical routine under high workload. It is likely that readers evaluated TOF-MRA scans more thoroughly than usual, as they were explicitly instructed to search for aneurysms. Third, the degree of automation bias might have been underestimated due to limited reader trust in the AI’s accuracy. This study featured an artificial sample with an unusually high rate of false-positive AI findings. Encountering questionable positive AI results in earlier cases might have led to diminished trust in the AI’s accuracy, influencing the ratings of subsequent cases. Future studies should evaluate automation bias in a larger, more representative cohort. Fourth, potential bias towards false-negative AI results was not assessed. With previously reported per-lesion sensitivities of aneurysm detection systems ranging widely from 67% to 100% (7), and given the high reliance of readers on the AI’s sensitivity in this study, it is conceivable that false-negative AI results lead to a higher frequency of missed aneurysms.

## Conclusion

Our results demonstrate vulnerability of radiology readers to automation bias in detecting cerebral aneurysms in TOF-MRA exams when faced with false-positive AI findings. Importantly, this behavior further translated into more aggressive follow-up recommendations among inexperienced readers. AI assistance resulted in significantly shorter reading times across experience levels, even in our artificial cohort with a very high rate of false-positive AI findings. While AI-assisted reading of radiology images can have benefits, challenges in human-AI interaction need to be mitigated to ensure safe and effective adoption.

## Data Availability

All data produced in the present study are available upon reasonable request to the authors

## Abbreviations

CAD: Computer-assisted diagnosis
CTA: Computed tomography angiography
TOF-MRA: Time-of-flight magnetic resonance angiography
DSA: Digital subtraction angiography
AI: Artificial intelligence

## References

1. Brown RD, Broderick JP. Unruptured intracranial aneurysms: epidemiology, natural history, management options, and familial screening. Lancet Neurol. Elsevier; 2014;13(4):393–404. doi: 10.1016/S1474-4422(14)70015-8.

2. Williams LN, Brown RD. Management of unruptured intracranial aneurysms. Neurol Clin Pract. American Academy of Neurology; 2013;3(2):99. doi: 10.1212/CPJ.0B013E31828D9F6B.

3. Menke J, Larsen J, Kallenberg K. Diagnosing cerebral aneurysms by computed tomographic angiography: Meta-analysis. Ann Neurol. John Wiley & Sons, Ltd; 2011;69(4):646–654. doi: 10.1002/ANA.22270.

4. Sailer AMH, Wagemans BAJM, Nelemans PJ, De Graaf R, Van Zwam WH. Diagnosing intracranial aneurysms with mr angiography: Systematic review and meta-analysis. Stroke. Lippincott Williams & WilkinsHagerstown, MD; 2014;45(1):119–126. doi: 10.1161/STROKEAHA.113.003133/-/DC1.

5. Park S, Lee DH, Ryu C-W, et al. Incidental Saccular Aneurysms on Head MR Angiography: 5 Years’ Experience at a Single Large-Volume Center. J Stroke. Korean Stroke Society; 2014;16(3):189. doi: 10.5853/JOS.2014.16.3.189.

6. Molenberg R, Aalbers MW, Metzemaekers JDM, et al. Clinical relevance of short-term follow-up of unruptured intracranial aneurysms. Neurosurg Focus. American Association of Neurological Surgeons; 2019;47(1):E7. doi: 10.3171/2019.4.FOCUS1995.

7. Din M, Agarwal S, Grzeda M, Wood DA, Modat M, Booth TC. Detection of cerebral aneurysms using artificial intelligence: a systematic review and meta-analysis. J Neurointerv Surg. British Medical Journal Publishing Group; 2023;15(3):262–271. doi: 10.1136/JNIS-2022-019456.

8. Shi Z, Miao C, Schoepf UJ, et al. A clinically applicable deep-learning model for detecting intracranial aneurysm in computed tomography angiography images. Nature Communications 2020 11:1. Nature Publishing Group; 2020;11(1):1–11. doi: 10.1038/s41467-020-19527-w.

9. Joo B, Ahn SS, Yoon PH, et al. A deep learning algorithm may automate intracranial aneurysm detection on MR angiography with high diagnostic performance. Eur Radiol. Eur Radiol; 2020;30(11):5785–5793. doi: 10.1007/S00330-020-06966-8.

10. Ueda D, Yamamoto A, Nishimori M, et al. Deep Learning for MR Angiography: Automated Detection of Cerebral Aneurysms. Radiology. Radiology; 2019;290(1):187–194. doi: 10.1148/RADIOL.2018180901.

11. Faron A, Sijben R, Teichert N, Freiherr J, Wiesmann M, Sichtermann T. Deep Learning-Based Detection of Intracranial Aneurysms in 3D TOF-MRA. AJNR Am J Neuroradiol. AJNR Am J Neuroradiol; 2019;40(1):25–32. doi: 10.3174/AJNR.A5911.

12. Lehnen NC, Haase R, Schmeel FC, et al. Automated Detection of Cerebral Aneurysms on TOF-MRA Using a Deep Learning Approach: An External Validation Study. American Journal of Neuroradiology. American Journal of Neuroradiology; 2022;43(12):1700– 1705. doi: 10.3174/AJNR.A7695.

13. Timmins KM, van der Schaaf IC, Bennink E, et al. Comparing methods of detecting and segmenting unruptured intracranial aneurysms on TOF-MRAS: The ADAM challenge. Neuroimage. Neuroimage; 2021;238. doi: 10.1016/J.NEUROIMAGE.2021.118216.

14. Sohn B, Park KY, Choi J, et al. Deep Learning-Based Software Improves Clinicians’ Detection Sensitivity of Aneurysms on Brain TOF-MRA. AJNR Am J Neuroradiol. AJNR Am J Neuroradiol; 2021;42(10):1769–1775. doi: 10.3174/AJNR.A7242.

15. Hirai T, Korogi Y, Arimura H, et al. Intracranial aneurysms at MR angiography: effect of computer-aided diagnosis on radiologists’ detection performance. Radiology. Radiology; 2005;237(2):605–610. doi: 10.1148/RADIOL.2372041734.

16. Miki S, Hayashi N, Masutani Y, et al. Computer-Assisted Detection of Cerebral Aneurysms in MR Angiography in a Routine Image-Reading Environment: Effects on Diagnosis by Radiologists. AJNR Am J Neuroradiol. AJNR Am J Neuroradiol; 2016;37(6):1038–1043. doi: 10.3174/AJNR.A4671.

17. Yang J, Xie M, Hu C, et al. Deep Learning for Detecting Cerebral Aneurysms with CT Angiography. Radiology. Radiology; 2021;298(1):155–163. doi: 10.1148/RADIOL.2020192154.

18. Lehnen NC, Schievelkamp A-H, Gronemann · Christian, et al. Impact of an AI software on the diagnostic performance and reading time for the detection of cerebral aneurysms on time of flight MR-angiography. Neuroradiology 2024. Springer; 2024;1–8. doi: 10.1007/S00234-024-03351-W.

19. Lyell D, Coiera E. Automation bias and verification complexity: a systematic review. Journal of the American Medical Informatics Association. Oxford Academic; 2017;24(2):423–431. doi: 10.1093/JAMIA/OCW105.

20. Khera R, Simon MA, Ross JS. Automation Bias and Assistive AI: Risk of Harm From AI-Driven Clinical Decision Support. JAMA. American Medical Association; 2023;330(23):2255–2257. doi: 10.1001/JAMA.2023.22557.

21. Agarwal Alex Moehring Pranav Rajpurkar Tobias Salz N, Lungren M, Langlotz C, et al. Combining Human Expertise with Artificial Intelligence: Experimental Evidence from Radiology. Cambridge, MA; 2023; doi: 10.3386/W31422.

22. Dratsch T, Chen X, Mehrizi MR, et al. Automation Bias in Mammography: The Impact of Artificial Intelligence BI-RADS Suggestions on Reader Performance. Radiology. Radiological Society of North America Inc.; 2023;307(4). doi: 10.1148/RADIOL.222176.

23. Goddard K, Roudsari A, Wyatt JC. Automation bias: A systematic review of frequency, effect mediators, and mitigators. Journal of the American Medical Informatics Association. Oxford Academic; 2012;19(1):121–127. doi: 10.1136/AMIAJNL-2011-000089/3/AMIAJNL-2011-000089FIG2.JPEG.

24. Lehnen NC, Haase R, Schmeel FC, et al. Automated Detection of Cerebral Aneurysms on TOF-MRA Using a Deep Learning Approach: An External Validation Study. AJNR Am J Neuroradiol. AJNR Am J Neuroradiol; 2022;43(12):1700–1705. doi: 10.3174/AJNR.A7695.

25. van Leeuwen KG, de Rooij M, Schalekamp S, van Ginneken B, Rutten MJCM. How does artificial intelligence in radiology improve efficiency and health outcomes? Pediatr Radiol. Springer; 2022;52(11):2087. doi: 10.1007/S00247-021-05114-8.

26. Neri E, de Souza N, Brady A, et al. What the radiologist should know about artificial intelligence – an ESR white paper. Insights Imaging. Springer Verlag; 2019;10(1):1–8. doi: 10.1186/S13244-019-0738-2/METRICS.

27. Brady AP, Allen B, Chong J, et al. Developing, purchasing, implementing and monitoring AI tools in radiology: practical considerations. A multi-society statement from the ACR, CAR, ESR, RANZCR & RSNA. Insights Imaging. Springer Science and Business Media Deutschland GmbH; 2024;15(1):1–19. doi: 10.1186/S13244-023-01541-3/FIGURES/1.

28. Mezrich JL, Mezrich JL. Is Artificial Intelligence (AI) a Pipe Dream? Why Legal Issues Present Significant Hurdles to AI Autonomy. 102214/AJR2127224. American Roentgen Ray Society; 2022;219(1):152–156. doi: 10.2214/AJR.21.27224.

29. Huisman M, Ranschaert E, Parker W, et al. An international survey on AI in radiology in 1041 radiologists and radiology residents part 2: expectations, hurdles to implementation, and education. Eur Radiol. Springer Science and Business Media Deutschland GmbH; 2021;31(11):8797–8806. doi: 10.1007/S00330-021-07782-4/TABLES/5.

30. van Leeuwen KG, Hedderich DM, Harvey H, Schalekamp S. How AI should be used in radiology: assessing ambiguity and completeness of intended use statements of commercial AI products. Insights Imaging. Insights Imaging; 2024;15(1). doi: 10.1186/S13244-024-01616-9.

31. Etminan N, de Sousa DA, Tiseo C, et al. European Stroke Organisation (ESO) guidelines on management of unruptured intracranial aneurysms. Eur Stroke J. Eur Stroke J; 2022;7(3):V. doi: 10.1177/23969873221099736.

32. Malhotra A, Wu X, Geng B, Hersey D, Gandhi D, Sanelli P. Management of Small Unruptured Intracranial Aneurysms: A Survey of Neuroradiologists. AJNR Am J Neuroradiol. AJNR Am J Neuroradiol; 2018;39(5):875–880. doi: 10.3174/AJNR.A5631.

