## Supplemental Table 1 for "Automation Bias in AI-Assisted Detection of Cerebral Aneurysms on Time-of-Flight MR-Angiography"

| case_id | finding_id | Age Range | Evaluation of AI Finding | Diameter of AI Finding (in mm) | Location | Description |
|---|---|---|---|---|---|---|
| **A01** | A01_1 | 41 – 45 | False Positive | 2.6 | Right M1 segment | Infundibulum |
| **A02** | A02_1 | 51 – 55 | True Positive | 2.8 | Left MCA bifurcation | Saccular aneurysm |
| **A03** | A03_1 | 51 – 55 | True Positive | 3.7 | Right MCA bifurcation | Saccular aneurysm |
| **A03** | A03_2 | 51 – 55 | False Positive | 1.7 | Left M1 segment | Infundibulum |
| **A04** | A04_2 | 56 – 60 | False Positive | 1.3 | Left M2 | Vascular loop |
| **A05** | A05_1 | 51 – 55 | True Positive | 4.6 | Right ICA | Saccular aneurysm |
| **A06** | A06_1 | 71 – 75 | False Positive | 2.8 | Right ICA | Infundibulum |
| **A07** | A07_1 | 66 – 70 | True Positive | 3.9 | Right ICA | Saccular aneurysm |
| **A08** | A08_1 | 81 – 85 | True Positive | 2.5 | Acom | Saccular aneurysm |
| **A09** | A09_1 | 76 - 80 | True Positive | 4.7 | Acom | Saccular aneurysm |
| **A09** | A09_2 | 76 – 80 | True Positive | 3.0 | Right MCA bifurcation | Saccular aneurysm |
| **A10** | A10_1 | 71 – 75 | False Positive | 2.9 | Acom | Acom fenestration |
| **B01** | B01_1 | 76 – 80 | False Positive | 1.4 | Right ICA | Vascular loop |
| **B02** | B02_1 | 16 – 20 | False Positive | 2.2 | Left M2 | Vascular loop |
| **B03** | B03_1 | 71 – 75 | True Positive | 5.6 | Left MCA bifurcation | Saccular aneurysm |
| **B03** | B03_2 | 71 – 75 | True Positive | 3.7 | Left ICA | Saccular aneurysm |
| **B04** | B04_1 | 56 – 60 | True Positive | 3.1 | Left MCA bifurcation | Fusiform aneurysm |
| **B05** | B05_1 | 31 – 35 | False Positive | 1.5 | Left M1 | Perforator |
| **B06** | B06_1 | 26 – 30 | False Positive | 2.0 | Left M1 | Perforator |
| **B07** | B07_1 | 31 – 35 | True Positive | 4.0 | Left ICA | Saccular aneurysm |
| **B08** | B08_1 | 26 – 30 | False Positive | 1.2 | Acom | Vascular loop |
| **B09** | B09_1 | 56 – 60 | True Positive | 4.0 | Left MCA bifurcation | Saccular aneurysm |
| **B10** | B10_1 | 31 – 35 | False Positive | 2.2 | Left MCA bifurcation | Vascular loop |

*Supplement 1: Overview of AI-positive findings.*
